# Exploring the Relationship Between Household Income and Healthcare Travel Patterns

**DOI:** 10.1101/2023.09.05.23295091

**Authors:** Amin Bemanian, Jonathan F Mosser

## Abstract

**Introduction:** Transportation is a social determinant of health which affects how easily individuals attend clinic appointments, obtain medications, or seek emergency care. Despite the importance of transportation, there are few large-scale studies characterizing typical transportation behaviors related to seeking healthcare. This study investigates demographic differences in travel times and transportation mode for healthcare/medical related trips using the National Household Travel Survey (NHTS) dataset and identifies potential access inequities.

**Methods:** Medical trip data was obtained from the 2017 NHTS dataset. All analysis was adjusted by survey weights. Multiple linear regression models were developed to investigate the relationships between demographic variables, transportation mode, and travel time. Additionally, a logistic regression model was used to investigate the relationship between demographics and transportation mode.

**Results:** Medical trips using public transportation were found to be significantly longer relative to personal automobile trips. Higher household income was found to be associated with shorter travel times, but the effect size decreased with the inclusion of transportation mode. Higher household income respondents were also likely to use public transportation. Respondent race was not significantly associated with any differences in travel times, but there were significant differences across races in public transportation use.

**Conclusions:** This study demonstrates that individuals with lower household incomes tended to have longer medical trips. Additionally, it identifies public transit to be a potential mediating factor for this difference, as lower-income residents were significantly more likely to use public transit. This analysis highlights the importance of maintaining accessible and efficient public transportation for access to healthcare, especially for populations which have traditionally been at-risk for health disparities.

## Introduction

Transportation to health services is often considered to be a determinant of health with transportation barriers and burdens often affecting healthcare access.^1–5^ Transportation barriers have been identified as a common reason for late, rescheduled, or missed healthcare appointments.^2,6–8^ One study by Wolfe et al. analyzing data from the National Health Interview Survey (NHIS) estimated that 5.8 million people in the United States delayed some form of medical care due to transportation barriers in 2017.^2^ Transportation barriers have also been associated with lower rates of medication and treatment adherence. A large retrospective study of pharmacy fillings for pediatric medications by Hensley et al. found that individuals living in ZIP codes with lower rates of vehicle ownership were less likely to successfully fill their prescriptions.^5^ Furthermore, transportation has been identified as affecting treatment adherence and efficacy for several chronic diseases including lung cancer chemotherapy,^9^ glycemic control in diabetes,^10^ antiepileptic usage,^11^ and antiretroviral therapy in HIV.^12^ These transportation barriers have been shown to disproportionately affect historically disadvantaged populations. Syed et al’s 2013 review on the relationship between transportation and health care access found five studies where transportation barriers or travel times were worse for racial or ethnic minorities compared to non-Hispanic Whites, including several which adjusted for the effect of socioeconomic status.^1^ In Wolfe et al.’s study of the 2017 NHIS data, there were significant differences in reporting transportation barriers across race/ethnicity, family income, employment, and educational attainment.^2^ There has been very limited research investigating the effects of transportation interventions for healthcare,^13^ but one study by Smith et al. investigated how expansion of a new light rail line affected missed medical appointments in Minneapolis and St. Paul, Minnesota.^14^ The study showed the creation of a new light rail resulted in a modest but significant decrease in missed treatments, with the effects being disproportionately concentrated among Medicaid patients.

While there is much literature that identifies potential and perceived relationships between transportation barriers and healthcare access, the majority do not actually measure the travel time or distance of health-related trips. Individual trip data has the benefit of allowing researchers and policymakers to ask more precise questions regarding the processes that cause these barriers such as the effect of transportation mode and vehicle ownership instead on relying on ecological data. One large dataset of individual trip data is the National Household Travel Survey (NHTS).^15^ The NHTS is a survey conducted by the Federal Highway Administration every 5 to 10 years with the goal of providing high resolution data for transportation agencies, urban planners, policymakers, and researchers. The survey logs individuals’ travel behaviors, including the purpose of each trip. There have been few prior health-related studies using this data. An analysis of the 2001 NHTS by Probst et al. found Black/African American participants were more likely to have to travel 30 minutes or more for a healthcare visit compared to White participants.^16^ Another study by Brucker and Rollins using the 2009 NHTS found that participants with medical conditions that limited mobility were more likely to have to travel 26 minutes or more compared to those without, despite not having significantly different trip lengths in miles.^17^ Currently, no studies have used the most recent version of the NHTS from 2017 to investigate the relationship between transportation and healthcare access.

### Objective

The purpose of this study is to characterize the distribution of medical trip travel times in the 2017 National Household Travel Survey across several different demographic variables and transportation modes in metropolitan and urban settings. Additionally, we test for population groups differences across these variables using multivariable models. Finally, we measure the relationship of transportation mode and travel times and test if there are any relationships between the demographic groups and the selection of transportation mode.

## Methods

### Data Source

The data source for this analysis was the 2017 NHTS.^15^ Households were invited via mail and participants were assigned a specific day to log all travel including trip duration (travel time), purpose, and mode of transportation. This information was then linked to the participant’s individual and household demographics. Households were then weighted to create a representative sample for the United States. For this analysis we included all trips whose purpose was listed as “medical/dental services” and whose participant was living in a metropolitan statistical area (MSA) at the time of the survey (n = 13,156 out of all 923,752 trips logged). Trips with travel times greater than 3 standard deviations from the mean were excluded as outliers (threshold of 102 minutes). Demographic variables of interest included participant age, sex, and race/ethnicity and household income, which were all collected using a self-reported survey at time of enrollment.

### Analysis

All analyses were done in R using the *survey* package to incorporate design weights in the analysis.^18^ Univariable and multivariable relationships with travel time were investigated using a series of linear multiple regression models. Due to the distribution of travel times, time was log-normalized. The first multivariable model (Model 1) included all participant demographic variables but excluded the transportation mode. The second model (Model 2) included all variables in the first and added transportation mode. We then stratified Model 1 by transportation mode to create Models 3a and 3b, which focused on personal automobile and public transit trips respectively. We then investigated the relationship between demographics and transportation mode. Univariable relationships were tested using a chi-squared test with Rao-Scott correction to adjust for design weights. Finally, a multiple logistic regression model (Model 4) was used to further investigate the relationship between demographics and transportation mode, with public transit usage as the outcome of interest. Using this combination of models, we tested whether transportation mode is a potential mediating pathway for the other relationships using the Baron and Kenny method.^19^ Mediation effects were tested using the Sobel test statistic.

## Results

Raw distributions of travel time stratified by respondent sex, race/ethnicity, and household income are shown in Figure 1. The survey-weighted mean travel time across all respondents was 23.6 minutes. The mean time among male respondents was slightly longer than female respondents (23.7 vs 23.6 minutes). American Indians/Alaska Natives (AI/AN) were found to have the longest mean time with 28.7 minutes while Asian Americans were found to have the shortest mean time of 19.8 minutes. For household income, respondents within the lowest household income stratum (<$25,000) were found to have the longest mean travel time of 28.0 minutes and within the highest income stratum (>$150,000) were found to have the shortest with 19.2 minutes. Public transit users had double the mean travel time relative to personal automobile users (45.0 minutes vs 22.2 minutes). We also calculated the survey-weighted contingency tables for the categorical variables, which are reported in the supplemental table S1.

**Figure 1.**
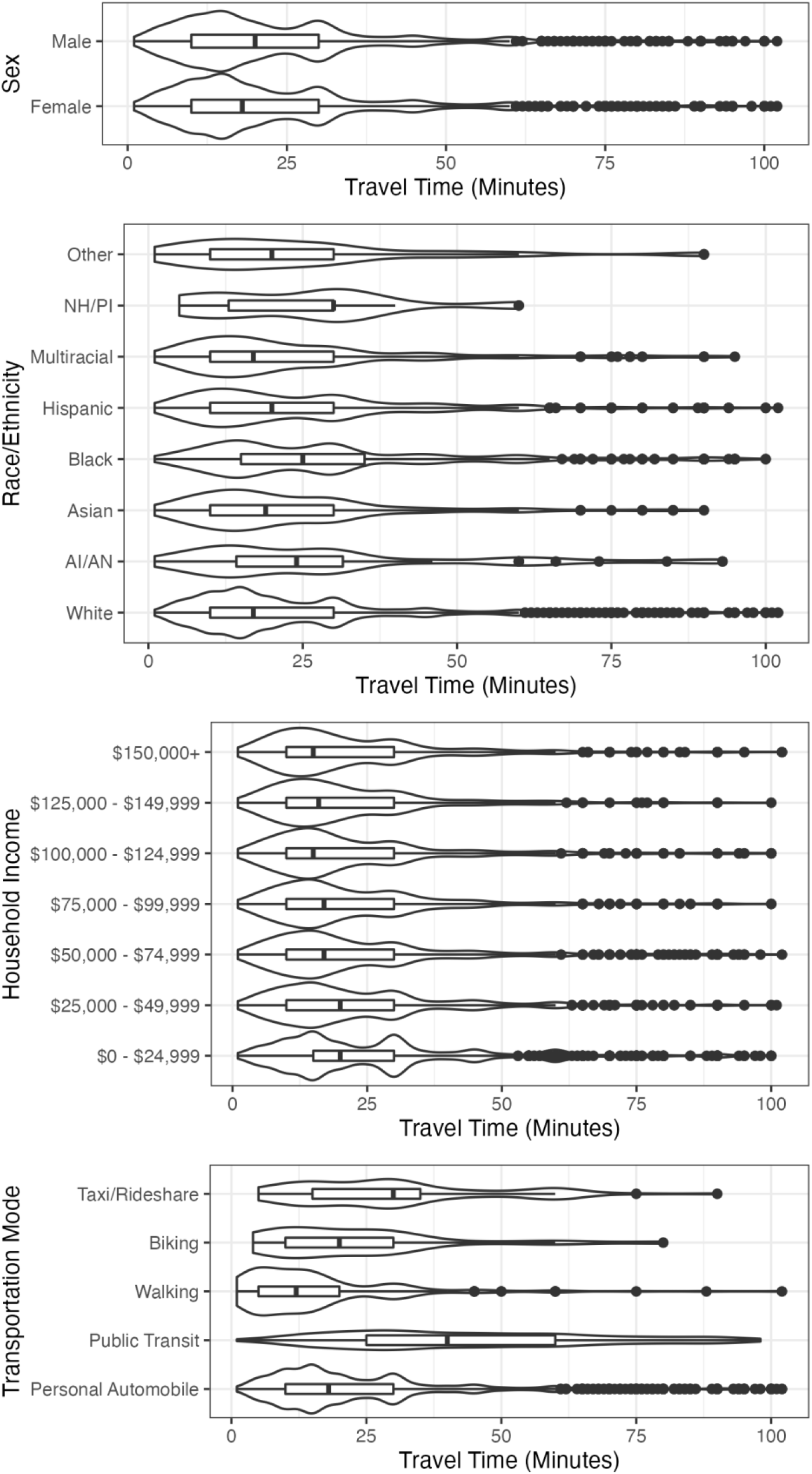
Violin plots showing the distribution of travel times across demographics and travel modes. Box-plot shows IQR and median for reference.

The first set of univariable models and multivariable models estimating travel time while controlling for demographic variables are shown in Table 1. In both Model 1 and the univariable models, when the lowest income stratum is used as a baseline, all other strata are found to have significantly shorter travel times. Additionally, older respondent age was associated with longer travel times. Respondent race/ethnicity and sex were not significantly associated with differences in travel times. In Model 2, after the addition of transportation mode the effect size of household income decreased across all strata and only two of the six strata still had statistically significant effect ($75,000 to $99,999 and > $149,000). Transportation mode was associated with differences in travel time with respondents who used public transit having significantly longer travel times and respondents who walked having significantly shorter travel times. There was no change to the effects of age, sex, and race/ethnicity.

**Table 1:** Model 1 and 2 looking at the relationship of demographic variables and transportation with travel times. All coefficients are reported as fold change with the 95% confidence interval in parentheses. Bolded parameters are significant at p < 0.05.

|  | *Univariable Models of*<br>*Travel Time w/*<br>*Demographics and*<br>*Transportation Mode* | *Model 1: Travel*<br>*Time w/*<br>*Demographics* | *Model 2: Travel*<br>*Time w/*<br>*Demographics and*<br>*Transportation Mode* |
| --- | --- | --- | --- |
| **Household Income** |  |  |  |
| <\$25,000 | 1 | 1 | 1 |
| \$25,000-49,999 | **0.850 (0.762, 0.948)** | **0.860 (0.771, 0.958)** | 0.952 (0.861, 1.054) |
| \$50,000-74,999 | **0.833 (0.760, 0.913)** | **0.850 (0.774, 0.933)** | 0.947 (0.866, 1.035) |
| \$75,000-99,999 | **0.788 (0.704, 0.882)** | **0.809 (0.723, 0.905)** | **0.888 (0.799, 0.988)** |
| \$100,000-124,999 | **0.817 (0.732, 0.913)** | **0.855 (0.760, 0.961)** | 0.957 (0.856, 1.070) |
| \$125,000-149,999 | **0.783 (0.682, 0.899)** | **0.835 (0.726, 0.959)** | 0.933 (0.813, 1.070) |
| > \$149,999 | **0.664 (0.588, 0.749)** | **0.701 (0.622, 0.791)** | **0.793 (0.710, 0.887)** |
| **Individual Age** | **1.005 (1.003, 1.007)** | **1.004 (1.002, 1.006)** | **1.004 (1.002, 1.005)** |
| **Sex** |  |  |  |
| Male | 1 | 1 | 1 |
| Female | 0.991 (0.926, 1.061) | 0.979 (0.916, 1.047) | 0.988 (0.928, 1.052) |
| **Race/Ethnicity** |  |  |  |
| White | 1 | 1 | 1 |
| American Indian/<br>Alaska Native | 1.067 (0.576, 1.976) | 0.984 (0.550, 1.760) | 0.809 (0.521, 1.257) |
| Asian | 0.885 (0.769, 1.020) | 0.922 (0.804, 1.056) | 0.928 (0.814, 1.057) |
| Black | 1.134 (0.992, 1.297) | 1.056 (0.929, 1.201) | 0.966 (0.855, 1.093) |
| Hispanic | 1.064 (0.958, 1.181) | 1.038 (0.936, 1.150) | 1.012 (0.919, 1.115) |
| Multiracial | 1.039 (0.917, 1.177) | 1.014 (0.896, 1.147) | 0.957 (0.856, 1.071) |
| Native Hawaiian/<br>Pacific Islander | 0.998 (0.678, 1.468) | 1.012 (0.691, 1.482) | 1.034 (0.700, 1.526) |
| Other | 1.128 (0.883, 1.442) | 1.128 (0.871, 1.461) | 1.144 (0.874, 1.496) |
| **Transportation Mode** |  |  |  |
| Personal Automobile | 1 | - | 1 |
| Public Transit | **2.155 (1.899, 2.446)** | - | **2.044 (1.796, 2.326)** |
| Walking | **0.543 (0.453, 0.650)** | - | **0.550 (0.465, 0.652)** |
| Biking | 0.559 (0.272, 1.147) | - | 0.609 (0.330, 1.124) |
| Taxi/Rideshare | 1.183 (0.889, 1.576) | - | 1.145 (0.869, 1.507) |

In Model 3a and 3b (Table 2), Model 1 was stratified by personal automobile and public transit use respectively. In Model 3a, only the highest income stratum (> $149,999) was significantly different from the lowest income stratum. All other variables were non-significant. In Model 3b, none of the variables were found to be significantly associated with differences in travel time. Table 3 shows the results of the logistic regression (Model 4) for using public transit versus other transportation modes. Higher household income strata were significantly associated with less public transit usage relative to the lowest stratum. In the survey-weighted contingency tables, 21.9% of the trips among the lowest income stratum were using public transit, while only 0 to 5% were using public transit for all the other income strata. Significant differences in race were also found for public transit usage. AI/AN, Black, Asian, Hispanic, and Multiracial respondents were found to have higher relative rates of public transit usage. Native Hawaiian/Pacific Islander and Other Race respondents did not have any occurrences of using public transit for medical trips, and their relative risk in the model was reported as 0. The Sobel test analysis showed significant mediation of public transit for the relationship between travel time and income across all income strata. Furthermore, significant mediation effects were found for race/ethnicity in AI/AN, Black, Hispanic, and Multiracial groups.

**Table 2:** Model 3a and 3b looking at the relationship of demographic variables on travel times stratified by transportation mode (personal automobiles and public transit). All coefficients are reported as fold change with the 95% confidence interval in parentheses. Bolded parameters are significant at p < 0.05. Note: There were no NH/PI or Other Race public transit users resulting in those coefficients to not be fit for model 3b. ^†^p-value of 0.0511, lower bound rounds up to 1.000.

|  | *Model 3a: Travel*<br>*Time for Personal*<br>*Automobile Users* | *Model 3b: Travel*<br>*Time for Public*<br>*Transit Users* |
| --- | --- | --- |
| **Household Income** |  |  |
| <\$25,000 | 1 | 1 |
| \$25,000-49,999 | 0.959 (0.859, 1.071) | 1.040 (0.726, 1.488) |
| \$50,000-74,999 | 0.940 (0.857, 1.031) | 1.208 (0.924, 1.579) |
| \$75,000-99,999 | 0.899 (0.802, 1.007) | 1.034 (0.654, 1.634) |
| \$100,000-124,999 | 0.979 (0.870, 1.101) | 0.837 (0.537, 1.305) |
| \$125,000-149,999 | 0.929 (0.806, 1.072) | 0.970 (0.545, 1.727) |
| > \$149,999 | **0.818 (0.727, 0.921)** | 1.279 (1.000, 1.637)<sup>†</sup> |
| **Individual Age** | **1.003 (1.002, 1.005)** | 1.000 (0.995, 1.006) |
| **Sex** |  |  |
| Male | 1 | 1 |
| Female | 1.001 (0.937, 1.070) | 0.922 (0.722, 1.178) |
| **Race/Ethnicity** |  |  |
| White | 1 | 1 |
| American Indian/Alaska Native | 0.688 (0.447, 1.059) | 1.243 (0.645, 2.396) |
| Asian | 0.882 (0.769, 1.012) | **1.564 (1.009, 2.424)** |
| Black | 0.968 (0.841, 1.115) | 1.091 (0.799, 1.490) |
| Hispanic | 1.017 (0.913, 1.132) | 1.139 (0.887, 1.464) |
| Multiracial | 0.946 (0.842, 1.064) | 1.040 (0.690, 1.569) |
| Native Hawaiian/Pacific Islander | 1.037 (0.704, 1.527) | - |
| Other | 1.161 (0.881, 1.531) | - |

**Table 3:** Model 4 looking at the relationship of demographic variables on the likelihood of taking public transit for a medical trip. Model coefficients are reported as the relative risk with the 95% confidence interval in parentheses. Sobel test statistic and p-value reported for the predicted mediation effect of each variable through public transit in the multivariate travel time model (Model 2). Note: There were no NH/PI or Other Race public transit users resulting in a RR of 0.

|  | *Model 4: Public*<br>*Transit Use vs Other*<br>*Modes* | *Sobel*<br>*Test*<br>*Statistic*<br>*for Travel*<br>*Time* | *Sobel*<br>*Test p-*<br>*value* |
| --- | --- | --- | --- |
| ***Household Income*** |  |  |  |
| <\$25,000 | 1 | | |
| \$25,000-49,999 | **0.163 (0.086, 0.310)** | **-4.936** | **< 0.001** |
| \$50,000-74,999 | **0.152 (0.082, 0.281)** | **-5.240** | **< 0.001** |
| \$75,000-99,999 | **0.239 (0.122, 0.468)** | **-3.898** | **< 0.001** |
| \$100,000-124,999 | **0.068 (0.022, 0.207)** | **-4.335** | **< 0.001** |
| \$125,000-149,999 | **0.038 (0.014, 0.101)** | **-5.601** | **< 0.001** |
| > \$149,999 | **0.108 (0.054, 0.218)** | **-5.399** | **< 0.001** |
| ***Individual Age*** | 1.009 (0.998, 1.020) | 1.618 | 0.106 |
| ***Sex*** |  |  |  |
| Male | 1 |  |  |
| Female | 0.972 (0.645, 1.463) | -0.137 | 0.891 |
| ***Race/Ethnicity*** |  |  |  |
| White | 1 |  |  |
| American Indian/Alaska Native | **9.050 (2.878, 28.457)** | **3.562** | **< 0.001** |
| Asian | 1.211 (0.433, 3.387) | 0.366 | 0.715 |
| Black | **3.936 (2.383, 6.503)** | **4.798** | **< 0.001** |
| Hispanic | **2.635 (1.576, 4.405)** | **3.500** | **< 0.001** |
| Multiracial | **3.089 (1.201, 7.944)** | **2.287** | **0.022** |
| Native Hawaiian/Pacific Islander | **0 (0, 0.00001)** | - | - |
| Other | **0 (0, 0.00001)** | - | - |

## Discussion

This analysis leveraged a large publicly available dataset to identify several important population differences in travel times and transportation modes for medical trips. Notably, lower income and older respondents had significantly longer travel times for these trips. Since travel time is intrinsically tied with transportation mode, it was important to determine the role mode played in these relationships. We found strong evidence for a relationship between household income and transportation mode, with the lowest income stratum being 4 to 25 times as likely to use public transit for health-related trips. Controlling transportation mode diminished the strength of the relationship between income and travel time, with the stratified analyses in Models 3a and 3b almost entirely removing the effect from Model 1. Transportation mode was a significant predictor of travel time, with public transit trips expected to be twice as long in duration relative to personal automobile trips. The combination of results across these models suggests that transportation mode likely mediates the relationship between household income and travel time.

Race/ethnicity was not found to be independently associated with travel time both in the univariable and multivariable models, despite prior studies finding evidence of a relationship.^1,2,16^ However, it is important to note there were racial/ethnic differences between the distributions of travel time, and race/ethnicity was a predictor of public transit use. Additionally, mediation analysis suggested a positive effect of race/ethnicity on transportation time via transportation mode. The significance of the mediation results for race/ethnicity is difficult to assess, however, since there were no independent effects identified in the univariable or multivariable models. Traditionally, the Baron & Kenny framework for mediation prescribes that there cannot be a clinically significant mediated relationship if there is no independent univariate association.^19^ However, other authors have argued this rule is too strict for non-experimental studies, and Strout & Bolger proposed an alternative framework where testing for an independent association is not necessary if the theoretical relationship is hypothesized to be “more distal,”^20^ This would include relationships which cannot be experimentally tested or if the theoretical relationship is expected to be relatively small and heavily mediated through other processes. Based on this alternate framework, there may be a significant relationship between race/ethnicity and travel time mediated by mode, especially given that we would expect any relationship between race/ethnicity and transportation time to be relatively distant and mediated through several processes. Additionally, these effects may be hard to measure in this study given the relatively small samples for non-Hispanic White individuals. Differences in general transportation mode usage and car ownership across race have been identified in previous studies, with non-Hispanic White individuals found to have higher rates of car ownership and lower rates of transit use compared to individuals of other races/ethnicities.^21,22^ Given the broader context of the health effects of transportation barriers, the differences identified in this study show how transportation can be a pathway for structural racism for health access and outcomes.

Older age was notably the only other significant predictor for longer travel times, which was also found to be a significant relationship in the Brucker and Rollins analysis of the 2009 NHTS but not in Probst et al.’s analysis of the 2001 NHTS.^16,17^ With mode stratification, the relationship between older age and longer time was unchanged among personal automobile trips but was not significant for public transit trips. Interestingly, there was no significant relationship between age and transportation mode, despite a 2017 study from Klein and Smart showing generational differences in car ownership.^21^

There are some important limitations with this study. First, while the NHTS does report a trip’s purpose as “medical/dental services”, this is self-reported by the participant and there is not specific instruction for what these trips should include. Therefore, it could include routine clinic visits, acute or emergent care, or long-term therapy visits. Additionally, there may be some classification discrepancies for pharmacy visits as the participant could list them as “shopping/errands” instead. Furthermore, there is a component of survivorship bias due to only actually completed trips being included in this dataset. We cannot account for healthcare trips that were delayed or canceled due to transportation barriers using this data. Finally, in our analysis, we chose only to include trips in metropolitan areas. While this limits the generalizability of this study’s results to rural settings, we felt including both would result in too much heterogeneity due to the fundamental differences in transportation behaviors and built environments in urban versus rural populations.

These findings highlight the disproportionate burden that households and individuals with lower incomes face from a transportation standpoint when trying to obtain medical care. Furthermore, it demonstrates why investment in affordable and efficient public transportation should be viewed as a public health need. Based on the findings of our analysis, further work is necessary to facilitate improved planning and coordination between healthcare organizations and municipal/regional transportation agencies to ensure all individuals have equitable physical access to healthcare.

## Data Availability

All data produced in the present study are available upon reasonable request to the authors and will be uploaded to a repository at a later date

---

**Supplement S1:** Contingency table of categorical demographic variables by transportation mode. Reported values are weighted counts instead of raw counts to adjust for survey weighting, Using a simulated sample pool of 1000 respondents. Note there may be some slight discrepancies due to rounding.

|  | Personal Automobile | Public Transit Use | Other | Total |
| --- | --- | --- | --- | --- |
| **Household Income** |  |  |  |  |
| Less than \$25,000 | 170.7 | 54.1 | 22.0 | 246.8 |
| \$25,000 - 49,999 | 211.0 | 8.7 | 6.7 | 226.4 |
| \$50,000 - 74,999 | 122.8 | 3.8 | 6.2 | 132.8 |
| \$75,000 - 99,999 | 97.1 | 4.7 | 3.5 | 105.3 |
| \$100,000 - 124,999 | 92.1 | 1.0 | 4.2 | 97.3 |
| \$125,000 - 149,999 | 55.7 | 0.3 | 1.8 | 57.8 |
| \$150,000 and higher | 121.5 | 2.3 | 9.7 | 133.5 |
| **Race/Ethnicity** |  |  |  |  |
| White | 568.5 | 21.0 | 25.8 | 615.3 |
| AI/AN | 2.0 | 1.0 | 0.0 | 2.9 |
| Asian | 30.9 | 1.2 | 2.9 | 34.9 |
| Black | 102.0 | 28.7 | 11.3 | 142.0 |
| Hispanic | 136.7 | 19.0 | 13.0 | 168.6 |
| Multiracial | 24.5 | 4.2 | 1.0 | 29.6 |
| NH/PI | 1.0 | 0.0 | 0.0 | 1.0 |
| Other | 5.5 | 0.0 | 0.1 | 5.5 |
| **Gender** |  |  |  |  |
| Female | 537.5 | 51.6 | 38.3 | 627.5 |
| Male | 333.4 | 23.4 | 15.7 | 372.5 |

## Notes

### Competing Interest Statement

The authors have declared no competing interest.

### Funding Statement

This study did not receive any external funding.

### Author Declarations

National Household Travel Survey NHTS 2017 Dataset: https://nhts.ornl.gov/downloads

